# Insights into the first wave of the COVID-19 pandemic in Bangladesh: Lessons learned from a high-risk country

**DOI:** 10.1101/2020.08.05.20168674

**Authors:** Md. Hasanul Banna Siam, Md. Mahbub Hasan, Md. Enayetur Raheem, Hasinur Rahaman Khan, Mahbubul H. Siddiqee, Mohammad Sorowar Hossain

**Affiliations:** Department of Emerging and Neglected Diseases, Biomedical Research Foundation, Dhaka, Bangladesh; Department of Genetic Engineering and Biotechnology, University of Chittagong, Chattogram 4331, Bangladesh; Department of Digital Health and Informatics, Biomedical Research Foundation, Dhaka, Bangladesh; Institute of Statistical Research and Training, University of Dhaka, Bangladesh; Department of Climate and Environmental Health, Biomedical Research Foundation, Dhaka, Bangladesh; Microbiology Program, Department of MNS, BRAC University, Dhaka, Bangladesh; School of Environment and Life Sciences, Independent University, Bangladesh (IUB), Bashundhara, Dhaka-1229, Bangladesh

**Keywords:** COVID-19, Pandemic, Bangladesh, Dhaka, Epidemiology, SARS-CoV2

## Abstract

**Background:** South Asian countries including Bangladesh have been struggling to control the COVID-19 pandemic despite imposing months of lockdown and other public health measures (as of June 30, 2020). In-depth epidemiological information from these countries is lacking. From the perspective of Bangladesh, this study aims to understand the epidemiological features and gaps in public health preparedness.

**Method:** This study used publicly available data (8 March-30 June 2020) from the respective health departments of Bangladesh and Johns Hopkins University Coronavirus Resource Centre. Descriptive statistics was used to report the incidence, case fatality rates (CFR), and trend analysis. Spatial distribution maps were created using ArcGIS Desktop. Infection dynamics were analyzed via SIR models.

**Findings:** In 66 days of nationwide lockdown and other public health efforts, a total of 47,153 cases and 650 deaths were reported. However, the incidence was increased by around 50% within a week after relaxing the lockdown. Males were disproportionately affected in terms of infections (71%) and deaths (77%) than females. The CFR for males was higher than females (1.38% vs 1.01%). Over 50% of infected cases were reported among young adults (20-40-year age group). Geospatial analysis between 7 June 2020 and 20 June 2020 showed that the incidences increased 4 to 10-fold in 12 administrative districts while it decreased in the epicenter. As compared to the EU and USA, trends of the cumulative incidence were slower in South Asia with lower mortality.

**Conclusion:** Our findings on gaps in public health preparedness and epidemiological characteristics would contribute to facilitating better public health decisions for managing current and future pandemics like COVID-19 in the settings of developing countries.

## 1 Introduction

The novel coronavirus disease 2019 (COVID-19) has triggered a public health emergency of international concern. Within a period of six months, the Severe Acute Respiratory Syndrome Coronavirus-2 (SARS-CoV-2) has spread to more than 200 countries/territories, infected more than ten million people worldwide, and claimed over half a million lives. The COVID-19 causes a plethora of clinical manifestations, and the severity and outcomes may vary depending on the underlying comorbidities (diabetes, heart diseases, hypertension, COPD), age, sex, and geographic locations (1). In South Asia, despite implementing various public health measures, the overall situation of the COVID-19 pandemic has been worsening and there is no sign of expected epidemic growth curve to be flattened. As of 12 July 2020, over one million cases and 31,688 deaths have been reported in the South Asia, and hence created an alarming situation as one-third of the world’s population (~1.7 billion) with similar socio-economic characteristics live in these densely populated and resource-limited regions. In this regard, Bangladesh could be an interesting setting to understand the characteristic of the COVID-19 pandemic from the South Asian perspective.

With a population of over 160 million, Bangladesh is one of the most densely populated (1265 per square km) countries in the world. About 60% of its population is between 15 to 64 years and only 4.7% is above 65 years of age (2). The care-home facility is virtually non-existent and the extended family structure combines the aged with the young in the same household, Because of economic prosperity, the country has been experiencing an increasing trend of unplanned urbanization, and currently, more than 32% of people live in urban areas. Bangladesh is also undergoing nutrition and epidemiologic transition with a higher burden of noncommunicable diseases (NCDs). A recent meta-analysis has shown that overall prevalence for metabolic syndrome (a cluster of health problems including high blood pressure, abdominal fat, high triglycerides, high blood sugar, and low HDL cholesterol) is higher in Bangladesh compared to the estimated world prevalence (30% versus 20-25%) (3). Besides, approximately 34% of adults are overweight and NCDs account for 67% of deaths in Bangladesh (4,5).

The capital Dhaka city has a population of nearly 20 million, and currently it is the epicenter of COVID-19 outbreak in Bangladesh. The first three official COVID-19 cases were reported on March 8, 2020, which included two men returning from Italy. Amid the upsurges of unofficial deaths of people with COVID-19 like symptoms, the first official death was confirmed on 18 March 2020. The city hosts more than 1 million slum dwellers and marginal communities who live in close proximity and are deprived of adequate facilities for maintaining personal hygiene as bathroom/toilets and water reservoirs are shared between several families (6,7).

Information on in-depth epidemiological features of the COVID-19 pandemic is lacking from the setting of developing countries, particularly from South Asia. In this context, we aim to (1) understand the gaps in public health preparedness, and (2) analyze the epidemiological characteristics including incidence, mortality, and geospatial distribution of the COVID-19 pandemic in Bangladesh. Moreover, comparative epidemiological trends were analyzed between Bangladesh and other countries with significant incidences of the disease.

## 2 Methods

### 2.1 Data sources

#### Incidence and mortality

Data were obtained from the Directorate General of Health Services (DGHS), the Government of Bangladesh (GoB) (8). Information on COVID-19 infected frontline professionals (medical doctor, health workers, police, and journalists) were extracted from the websites of Bangladesh Medical Association, major newspapers and online portals, professional societies and then verified with the government press releases whenever possible (9-13).

Unofficial death counts with COVID-like symptoms were collected from weekly bulletins published by Bangladesh Peace Observatory, Centre for Genocide Studies (CGS) at the University of Dhaka along with the official death counts (14).

#### Geospatial analysis

District-wise case reports data was collected from the Institute of Epidemiology, Disease Control and Research (IEDCR), a sister organization of DGHS and COVID-19 Bangladesh situation reports by World Health Organization (WHO) (15,16). District-wise population data were retrieved from the Bangladesh Population Census 2011 dataset. The spatial map was created using layers downloaded from The Bangladesh Geospatial Data Sharing Platform (GeoDASH) on ArcGIS Desktop (Esri Inc., Redlands, California, United States) (17).

#### Trends analysis and modeling

Data from the Johns Hopkins University Coronavirus Resource Centre was used to analyze the current trend of Bangladesh compared to South Asian countries (India, Pakistan, and Nepal) and other countries with higher incidence (Saudi Arabia, Brazil, UK, USA, and Italy) (18). For SIR models (Bangladesh, India, Pakistan, and Nepal) COVID-19 data from the above-mentioned source and population data were obtained from Our World in Data.

### 2.2 Statistical analysis

Descriptive statistics were used to report incidence, mortality, age, and gender-specific attributes. To model the COVID-19 infection dynamics, a simple SIR model was used, which is particularly suitable to deal with a large population. The initial model considers the N number of populations to be classified into three categories: the susceptible (S), the infected (I), and the recovered (R). The rate of transition from susceptible to infected is denoted by *β* or the transmission rate; and the rate of transition from infected to recovered is by *γ* or the recovery rate. We assumed that the population in our study was closed and individuals mixed uniformly in the community. Following Britton and Giardina, we also assumed that all individuals were equally susceptible to the disease and equally infectious if they get infected (19). We fitted the SIR model for each country considered in this study. The current total population for each country was taken from Worldometer. We used L-Broyden–Fletcher–Goldfarb–Shanno algorithm (L-BFGS) to optimize the *β* and *γ* parameters of the SIR model while 0.5 was considered as the initial value for each of the parameters. The L-BFGS was a limited-memory version of BFGS and generated the optimized *β* and *γ* parameters using the COVID-19 observed incidences. The following equations were used in the SIR model:

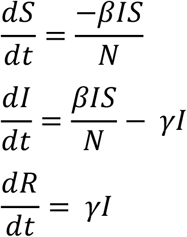

## 3 Result

### 3.1 Major events, the trend of infection and deaths in the first four months of COVID-19 outbreak

Figure1 represents an overall timeline of COVID-19 related major events in the first four months in Bangladesh. Bangladesh officially started imposing intervention measures to tackle COVID-19 in March 2020. After the first case was reported, flights were limited, educational institutions were closed, and health screening was imposed at all the port of entries. Two weeks after the first confirmed case, the government imposed a nation-wide lockdown (ref). During this period, all offices remain closed, public transportation was shut, gatherings and all sorts of inter-district travels were suspended, prayers in places of worship were suspended, and mandatory stay at home order was imposed. The lockdown was relaxed after 66 days at which point the total COVID-19 cases reached 47,153 with a death toll of 650. Within a week, reporting of new cases increased by roughly 50%. After three months into the pandemic, the total cases reached 68,508; among them, 14,560 (21.25%) were recovered and 930 (1.36%) were deceased.

**Figure 1:**
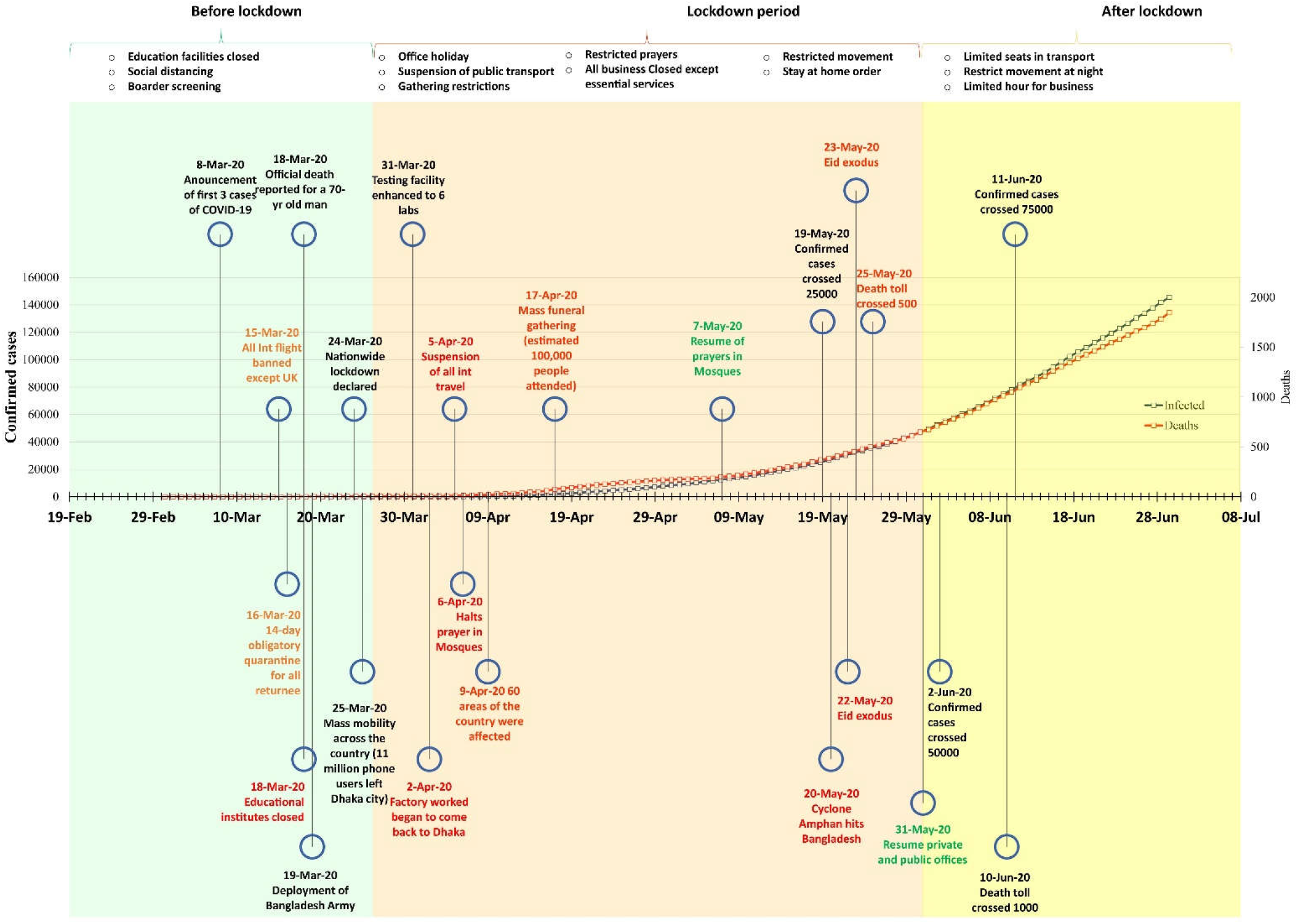
Major events and public health measures in COVID-19 outbreak in Bangladesh. Deaths over time are represented in the secondary axis.

### 3.2 Demographic characteristics of COVID-19 cases and mortality

More than 50% of patients infected with SARS-CoV-2 were aged between 21 and 40 years (Figure 2A). Children and elderly people belong to the least infected group. Deaths due to COVID-19 in Bangladesh increased with the age of patients (Figure 2B). About 40% of cases aged over 60 years were four times more likely to die than the age group of 31-40 years. Males were infected (71%) and died (77%) in a higher proportion than females. (Figure 2C-D). Overall case fatality rate (CFR) in Bangladesh was 1.27% while CFR for males was higher than females (1.38% vs 1.01%).

**Figure 2:**
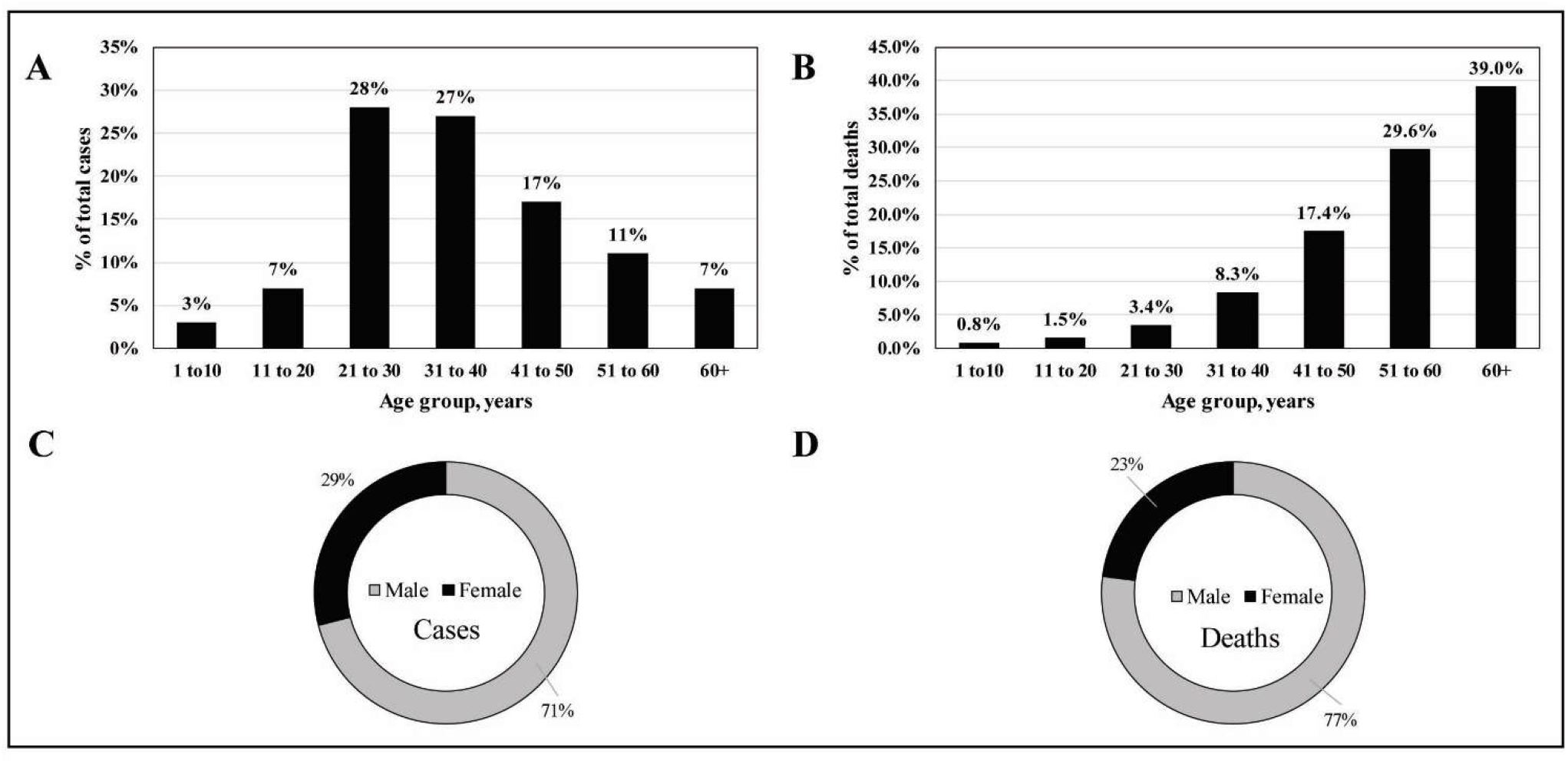
Demography of reported COVID-19 cases and deaths in Bangladesh (8 march 2020-30 June 2020). A) Age group specific cases B) Age group specific deaths C) Sex specific cases D) Sex specific deaths.

As of 12 July 2020, over 20,000 frontline health workers (5483), law enforcement officers, and journalists were infected (Table 1). A high CFR (3.34%) was recorded for frontline physicians. A total of 1500 unofficial deaths (as of 2 July 2020) with COVID-19 like symptoms were reported (Figure 3). In the first three weeks of April, the unofficial death of COVID-19 suspects increased rapidly in the Dhaka division while deaths increased alarmingly in Chattogram division after June 7, 2020, and now most deaths reported from this division with a death toll of 451. The trend of an increase in the number of deaths was slow in Barishal, Rangpur, Mymensingh, Sylhet, Khulna, and Rajshahi as compared to Dhaka and Chattogram.

**Figure 3:**
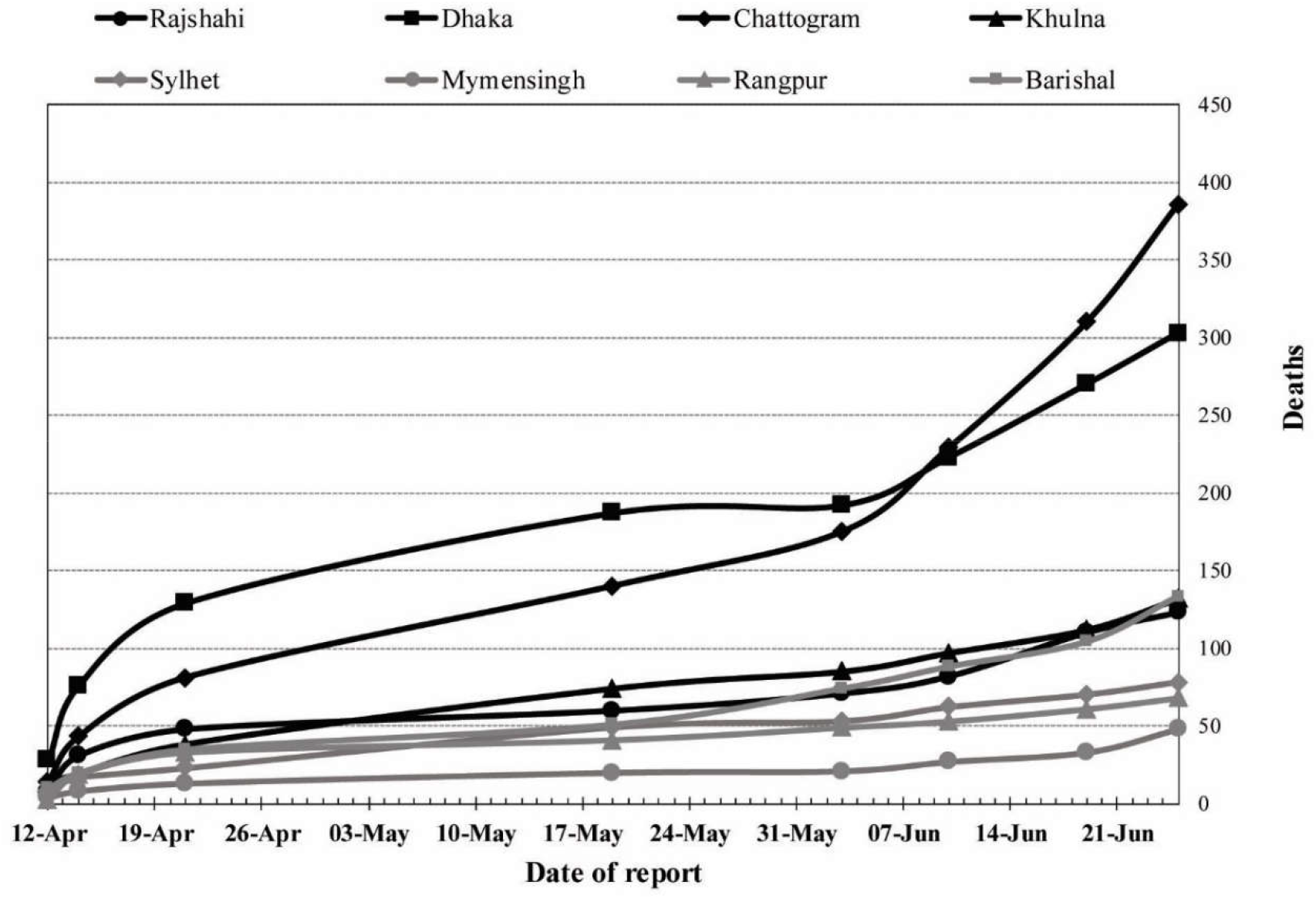
Trends of unofficial deaths with COVID-19 like symptoms in different divisions across Bangladesh

**Table 1:** Infections and Deaths among frontline COVID-19 fighters (Police, health workers, Army, Journalists)

| Frontliners | Infected (n) | Death n (%) |
| --- | --- | --- |
| Physicians | 1886 | 63 (3.34%) |
| Nurse | 1502 | NA |
| Medical Staff | 2095 | NA |
| Police | 11302 | 44 (0.38%) |
| Army** | 3477 | NA |
| Journalists | 378 | 06 (1.58%) |

\*\*Both in service and retired

The testing facility was minimal in the first month of the outbreak (March 2020) and later increased to 66 laboratories nationwide. Albeit the overall testing rate was still only 4.7 tests per 1000 people and was below the rate of India, Pakistan, Sri Lanka, and other South Asian countries (Suppl. Figure 1).

### 3.3 The district-wise burden of COVID-19 in Bangladesh

District-wise data for active COVID-19 cases (excluding recovered and deceased cases) were retrieved on two-time points (June 7, 2020, and June 30, 2020). The infection rate rose to 521 cases per million on June 30 from 275 cases per million on June 7^th^, nearly a 90% increase over three weeks (Figure 4). Dhaka had the highest incidence rates at both time-points but the number decreased to 1507 from 1735 cases per million (13% reduction). This may suggest that the disease spread over the country except for the epicenter Dhaka. The rapid spread of the disease was observed in Tangail (from 15 cases per million to 168 cases) and Khulna (from 66 cases to 742 cases per million). That is more than a 10-fold increase over three weeks. Twelve districts out of 64 reported a 4 to 10-fold increase of COVID-19 incidences.

**Figure 4:**
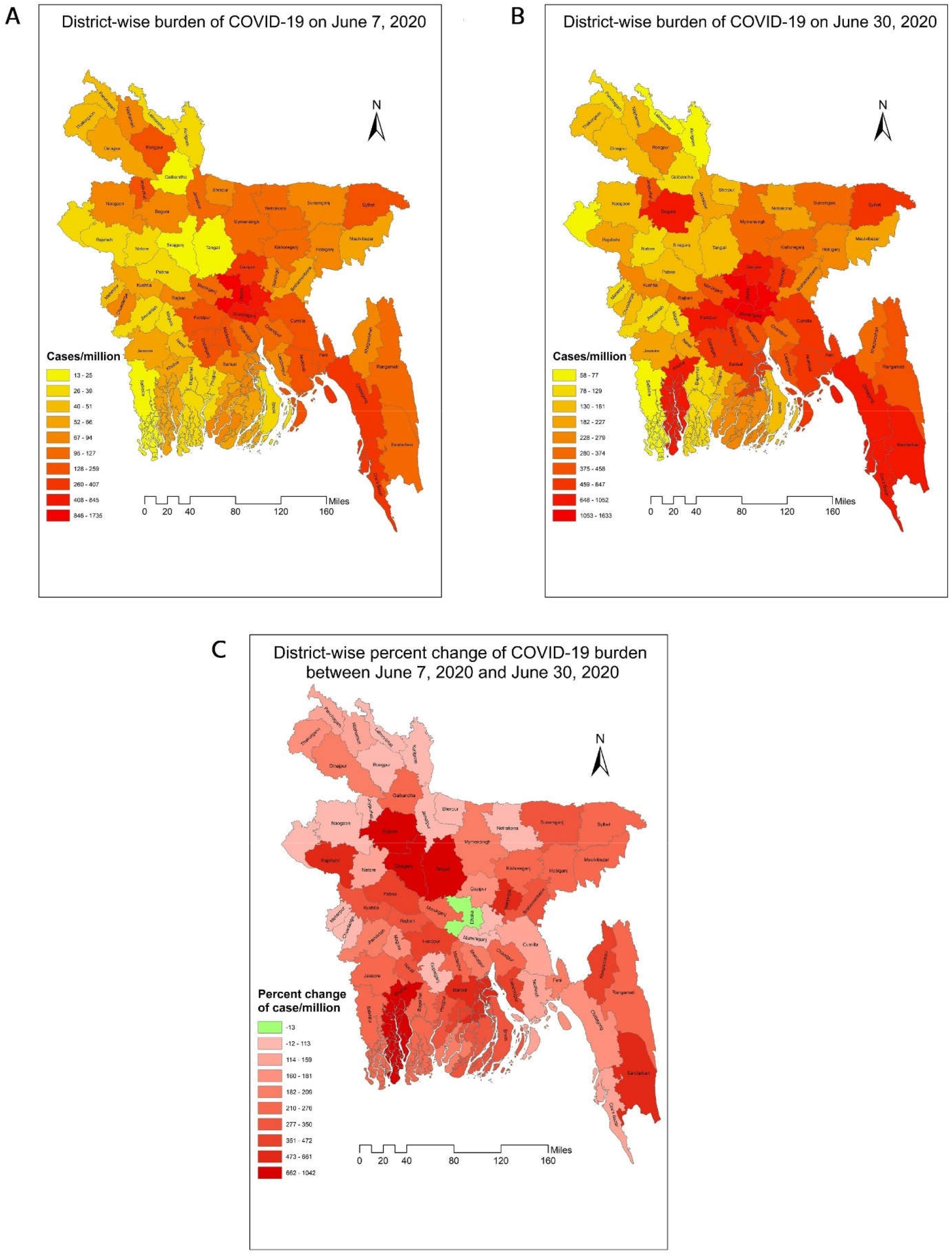
Geospatial distribution of COVID-19 in Bangladesh. A) district-wise distribution of active COVID-19 cases on June 7, 2020 B) district-wise distribution of active COVID-19 cases on June 30, 2020 C) district-wise percent change of active COVID-19 cases between June 7, 2020 and June 30, 2020

### 3.4 Trends of cumulative incidence and mortality of Bangladesh compared to other countries

The trends of incidence and mortality in Bangladesh were compared with some South Asian and other countries with significant COVID-19 infections. In Figure 5, we found that Bangladesh had a similar trend of incidence during the initial phase of the pandemic (first 30 days or so). After that, the incidence in Bangladesh and other South Asian countries slowed down compared to Brazil, the USA, and European countries. As for mortality, the South Asian countries had significantly fewer deaths over time compared to the other countries in the EU and the USA. Interestingly, both incidence and deaths in Nepal started to pick up after more than one and a half months since the first confirmed case.

**Figure 5:**
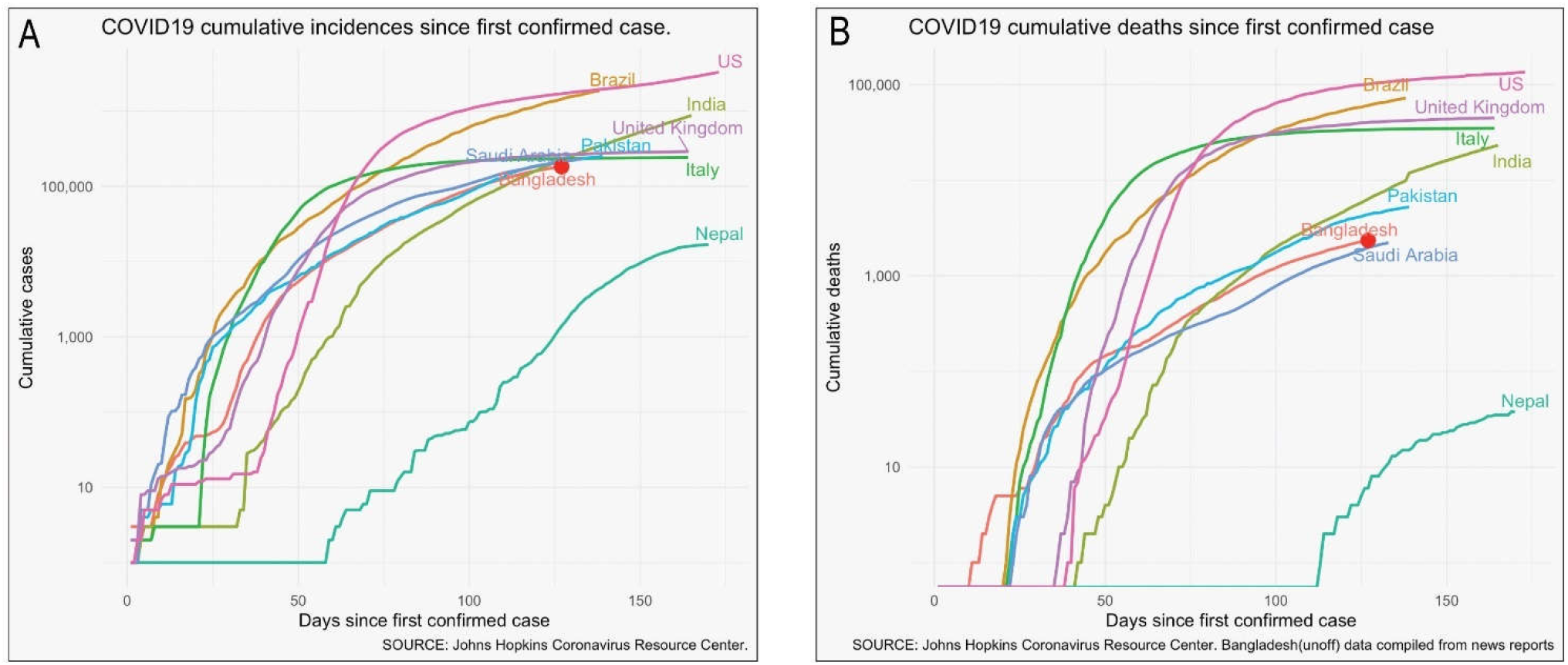
Trends of COVID-19 cumulative incidences and deaths in Bangladesh as compared to other South Asian and Western countries.

### 3.5 Infection dynamics via SIR models

Given predictions obtained by the SIR model till June 28, with the same settings, Bangladesh was expected to reach the peak by the end of July (Table 2, Figure 6). About 852,525 people would be infected by then, which translates to about 170,505 severe cases who would need hospital admissions and over 34,100 persons in need of intensive care and up to 10,753 deaths (assuming a 1.26% fatality rate, as found in Bangladesh as of 28 June 2020. The peak in India, Pakistan, and Nepal were expected to be reached on July 23, August 10, and August 5 respectively (Table 2).

**Figure 6:**
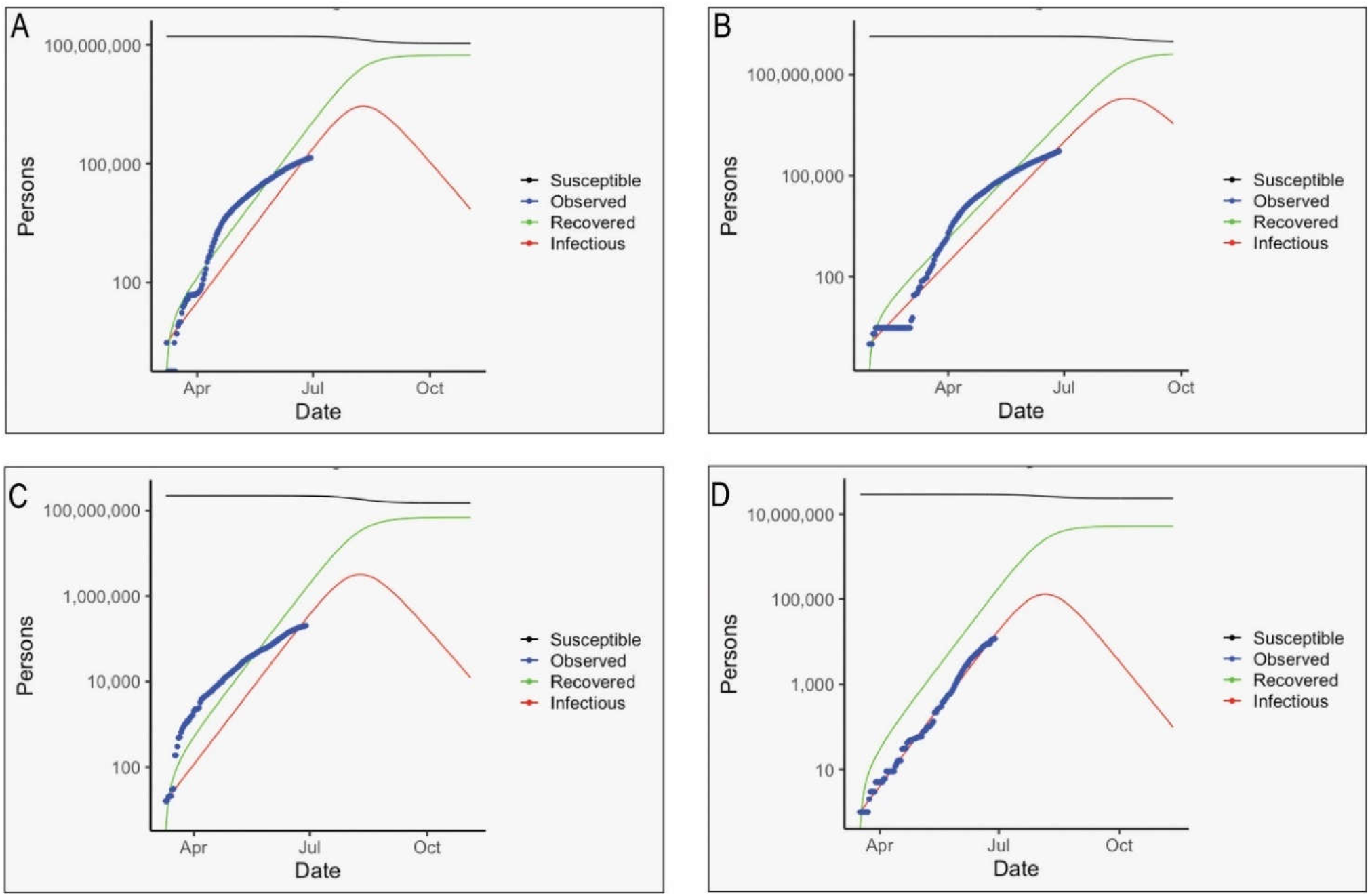
SIR models fitting for COVID-19 incidence in selected countries. A) Bangladesh B)India c) Pakistan and D) Napal

**Table 2:**
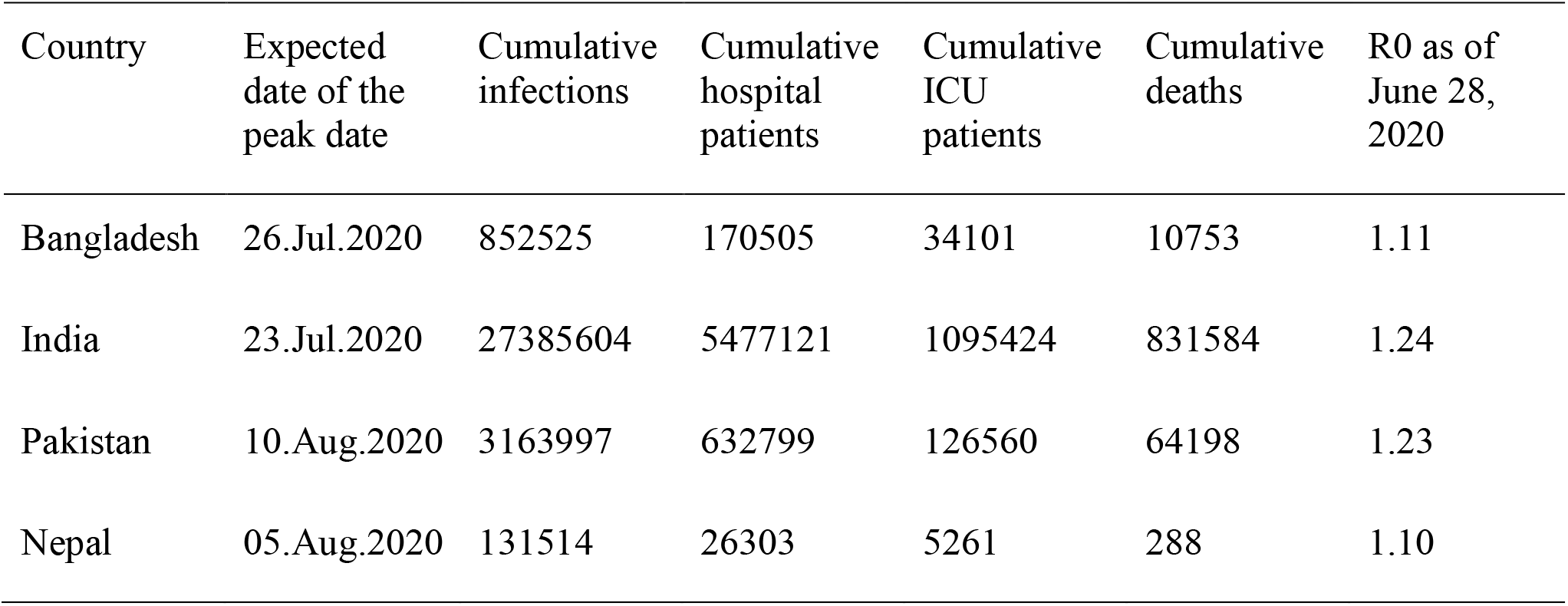
Expected Peak date, COVID-19 incidence, and health capacities of Bangladesh as compared to other South Asian countries

## 4. Discussion

The COVID-19 pandemic has appeared as a daunting disaster for Bangladesh as the country now transitions towards its highest peak in the number of positive cases. This article is the first attempt to describe the public health preparedness and epidemiological characteristics of the first wave of COVID-19 pandemic in Bangladesh.

### 4.1 Ineffective public health crisis communication and countermeasures

After the official announcement of COVID-19 cases in Dhaka, the Government took a number of initiatives to revamp the testing facility, medical equipment supply, and nationwide surveillance. The lockdown measures with the deployment of police and armed forces were routed to enforce the law. However, our study shows that public health countermeasures were not up to the mark to flatten the curve as the country has been witnessing a steep rise in the number of confirmed cases. A series of events such as the mobilization of 11 million mobile users from city to rural areas, local violence, massive gathering for Janaza prayer (funeral), and purposive rule-breaking indicated that people, by and large, failed to take into account the importance of maintaining social distancing. While strict lockdown policy has shown success in China, New Zealand, Italy, and largely in the developed countries, a similar approach did not achieve success in Bangladesh, for which the underlying socioeconomic and demographic factors were at fault. At the beginning of April during the lockdown period, Bangladesh had only 54 cases, but it increased to about 50,000 cases by the end of May as the lockdown ended. A similar trend was also observed in India and Pakistan where the early lockdown measures might have helped in the preparation of medical logistics but ultimately failed to contain the viral spread.

One recent study in Bangladesh has shown that people were confused about the "english" terms such as “stay at home,” “social distancing,” “quarantine,” and “lockdown” (20). The upward trajectory of positive cases despite an extended period of nationwide lockdown and restrictions indicate the inefficiency of public health risk communication. The gap in the public health preparedness in battling COVID-19 can be minimized by taking into account the nuances of geographical & cultural contexts and devising specific strategies. Significant investment and support are required to generate sufficient data for improving the efficiency of public health decisions.

### 4.2 Gender disparity and spatial distribution of infection

In Bangladesh, the proportion of men catching the virus was 2.5 times more than women, although studies around the world suggest that both men and women are equally susceptible to the virus (15,21). A similar observation was noted in India and Pakistan where men comprised about 65% and 70% of confirmed cases, respectively (22,23). This disparity in susceptibility might be due to cultural aspects since men dominate the outdoor activities and are less careful towards keeping up with personal hygiene. Also, the percentage of susceptibility might vary since COVID-19 is more fatal for men, causing them to seek medical care and subject to testing (24). The death rate among males was found higher in various studies around the world. In South Asia, males died in a higher proportion: India (64%), Pakistan (74%), and Bangladesh (77%). Studies showed that men over 60 years of age are twice as likely to die of COVID-19 than women (24,25). This could partly be explained by the presence of higher comorbidity and the smoking habit in men, although the exact reason remains to be elucidated (26). However, women in India had a higher case fatality rate compared to males (CFR: 2.9% versus 3.3%) which contrasts the scenario of Bangladesh (CFR: 1.4% versus 1.01%) (21,22). Future studies are warranted to understand the disparity in sex-specific mortality risk in South Asia.

In Bangladesh, the existing data suggest that the infection rate in people from urban areas is higher than in rural villages. This is unsurprising because urban settings offer a higher chance of catching the virus. For instance, the Dhaka or Chittagong city relies heavily on congested infrastructures where the population density is the highest, and the citizens receive reduced sunlight exposure. Vitamin D deficiency in city areas is well documented, and some early studies indicated that vitamin D deficiency could be a risk factor for COVID-19 adversity (27,28).

On the flip side, the rural remote regions of the country suffer from a lack of proper medical support. Out of 64 districts, almost half of the districts have over 100,000 people who are aged above 65 years, and data show that the proportion of the elderly is higher in rural areas (29,30). As hospital facilities and medical assistance are more centered towards urban residents, about 3 million people from 50 tribal and ethnic minor communities living in remote areas remain vulnerable to COVID-19 (29). In Brazil, around 9.1% of indigenous people infected with the disease died, and the rate of infection is soaring in distant communities (31). Correspondingly, the Rohingya refugee forcibly displaced from Myanmar are living in crowded quarters in Cox’s Bazar area, Bangladesh, where they are subject to several health issues such as malnutrition, limited medical access, food- and water-borne diseases, reproductive health, and communicable diseases. In addition to Bangladesh Government’s support, the UN initiative to mobilize women in spreading awareness regarding COVID-19 in Rohingya camps might lessen the disease burden; however, additional international aid is necessary to cope up with the challenges (32).

### 4.3 Low fatality rate as compared to other countries

Interestingly, despite the official numbers jumping above 186,000 (as of 12 July 2020) cases, the death rate remained fairly low (1.27%) in Bangladesh and close to the other South Asian countries such as Afghanistan (2.93%), India (2.64%) and Pakistan (2.09%). This trend contrasts strongly to the other countries such as France (14.5%), Italy (14.3%), the UK (13.9%), Spain (10.4%), USA (4.07%), China (5.54%), Japan (4.39%) and Iran (4.99%) (33). The lower mortality rate in the developing countries may be due to the fact that the overall life expectancy is within 64 to 72 years, leaving out the overly elder population; and there is a higher proportion of young populations in Afghanistan, Pakistan, India, and Bangladesh. By comparison, the proportion of people aged over 70 years is much higher in the European and American settings (34).

Speculations regarding the low death rate in developing countries questioned the under-reporting of actual death cases as testing facilities were very limited. The unofficial sources indicated that a slightly higher number of deaths (n=1500) occurred in Bangladesh during the initial phase of the outbreak. Over the last four months, the Bangladesh Government has increased its testing capacity nationwide, yet the combined overall mortality rate remains surprisingly lower than what has been observed in the western world. Importantly, the western countries offer care home facilities which are practically nonexistent in South Asian culture where the extended family structure incorporates the elderly with the young in the same household. As opposed, the rapid spread of the infection among the elderly in care homes contributed to half of the total deaths in the western world. In France, the deaths linked to care homes were 51%, and in Canada, as many as 82% of COVID-19 deaths occurred among care home residents (35).

### 4.4 Slow progression of COVID-19 pandemic in South Asian countries

Despite higher population density and low awareness of personal hygiene, the South Asian countries have witnessed a rather noticeable slow progression. Our model also demonstrated a similar scenario where the peak infection is coming much later than any of the western countries although South Asian countries are closer to and have strong economic relations with China. Of note, the first COVID-19 positive case was identified on January 30 in India, February 26 in Pakistan, and March 8 in Bangladesh. Despite such early detection, the subsequent numbers of infection and mortality continued to rise slowly. Several factors might influence the disease progression such as the wide-scale adaptation of facemask, tropical climate, or cross-immunity from other viruses as the region is under the high burden of disease (36,37). However, the most probable reason is that, unlike China that locked down Wuhan city, the South Asian countries went for a nationwide total lockdown. Such actions had a tremendous impact on the daily lives of people, especially the daily wage earners but the measures turned out to be unsustainable. Now that countries are easing down on lockdown measures, the numbers are soaring up. In Bangladesh, the post-lockdown COVID-19 infection is dispersing quickly from the cities towards the peripheral districts, and a dramatic shift in the percent-change of disease transmission is observed. This underpins the fact that relaxing social distancing measures might bring about similar consequences for developing countries.

### 4.5 Overburden of healthcare system: higher fatality rate among medical doctors, limited testing, and ICU beds

A significant number of medical personnel and law enforcement workers were infected during the first wave of the COVID-19 epidemic. In particular, the fatality rate among the doctors in Bangladesh (3.34%) was higher than the healthcare workers in Pakistan (1.01%) and India (3.30%) and slightly lower than Afghanistan (3.75%) (38). The higher proportion of frontliners contracting the virus implies the lack of personal safety and inconsistency in management. The quality of personal protective equipment (PPE) and its training of proper handling remains in doubt. Moreover, hospitals suffer from the scarcity of intensive care units. With only 733 ICU beds in government hospitals, the healthcare has crumbled as COVID-19 infection rate increases in torrent (39). It is surmised that the number of deaths in the age range of 60–70 could be subverted if proper ICU and medical support were provided. Our model implied that the number of hospitalized and ICU patients will drastically increase during the peak period of infection which will overload the medical capacity. This underscores the need for urgent investment and remodeling of the healthcare sector.

Undoubtedly, the biggest problem with social distancing in Bangladesh lies in the population density and city-based centralized facilities. Besides, the healthcare system in Bangladesh is also largely centralized in metropolitan settings especially capital Dhaka and divisional cities. The disaster of COVID-19 pandemic thus points out the fragility of the existing system and urges to prioritize the decentralization of healthcare facilities. The steps can also ensure the minimization of the urban-rural disparity in the healthcare facilities. The establishment of universal health care should also be directed as a long-term goal.

The abrupt onset of critical patients at the hospitals has strained medical resources; and expensive PCR based testing facilities and related workforce remained in pestilence to meet the demand. Despite the upscaling of testing labs, Bangladesh is lagging behind the race in regard to tests per 1000 people. Interestingly, Japan has tested fewer than Bangladesh and still maintains a very low infection and mortality rate. This indicates that proper contact tracing and quarantining is equally important. Since public health measures in the developing countries are constrained by resources both in terms of financial support and trained workforce, it is essential to devise a strategy that is most suitable for such settings. It is recommended that instead of depending on RT-PCR based tests that are costly and time-consuming, the low-middle income countries could focus on syndromic diagnosis based on the constellation of symptoms and signs of COVID-19 in a disease afflicted area (40). This way, the burden of expensive testing could be largely reduced, the suspected patients identified earlier, and the investment channeled out into rebuilding healthcare facilities. Moreover, patients who were denied hospital admittance because of the absence of COVID-19 test report will also receive early medical support.

In the last four months, the Bangladesh government has scaled up the testing facility across the country as 66 labs are now providing RT-PCR based COVID-19 detection tests. Although the situation is not going to be under control any time soon, by the time COVID-19 ends, the testing labs should be converted into regional surveillance centers instead of dissolving them. A nationwide sustainable monitoring and coordination system will help in swift action in case of future outbreaks.

Our study has a few limitations. First, we used publicly available recent data for analysis, which was both a weakness and a strength in itself. Second, for comparative infection dynamics, SIR model was developed on the assumption that lockdown and social distancing did not restrict the movements of people, therefore people had an equal chance of contact with every other person among the population irrespective of space or distance. However, this assumption might not reflect the real situation uniformly in all countries. Third, unofficial death counts with COVID-19 like symptoms (not confirmed by the test) were extracted from well-established national news media and thus, these should be considered as probable death cases. But, Due to limited healthcare infrastructure and testing facilities, it was not possible to confirm all suspected deaths.

## Conclusion

We found that despite imposing a number of public health measures to ‘flatten the curve; the outbreak continued to grow and spread across Bangladesh. Males were disproportionately infected and died at a higher percentage. Case fatality rates were relatively higher among frontline medical workers and law enforcement officials. South Asian countries including Bangladesh had significantly fewer deaths over time compared to the other western countries. Our finding on gaps in public health preparedness and communication, and epidemiological characteristics would contribute to better public health decisions for managing current and future pandemic like COVID-19 in Bangladesh and other similar settings elsewhere in the world, particularly in South Asia.

## Data Availability

Available

## Conflict of Interest

The authors declare that the research was conducted in the absence of any commercial or financial relationships that could be construed as a potential conflict of interest.

## Author Contributions

MSH conceived the idea. MSH, MMH and MHBS designed and prepared the first draft. MMH, MHRK, ER and MHS contributed to the result preparation. All authors critically reviewed and edited the manuscript.

## Funding

No funding was associated with this work.

## Supplementary materials

**Supplementary Figure 1:**
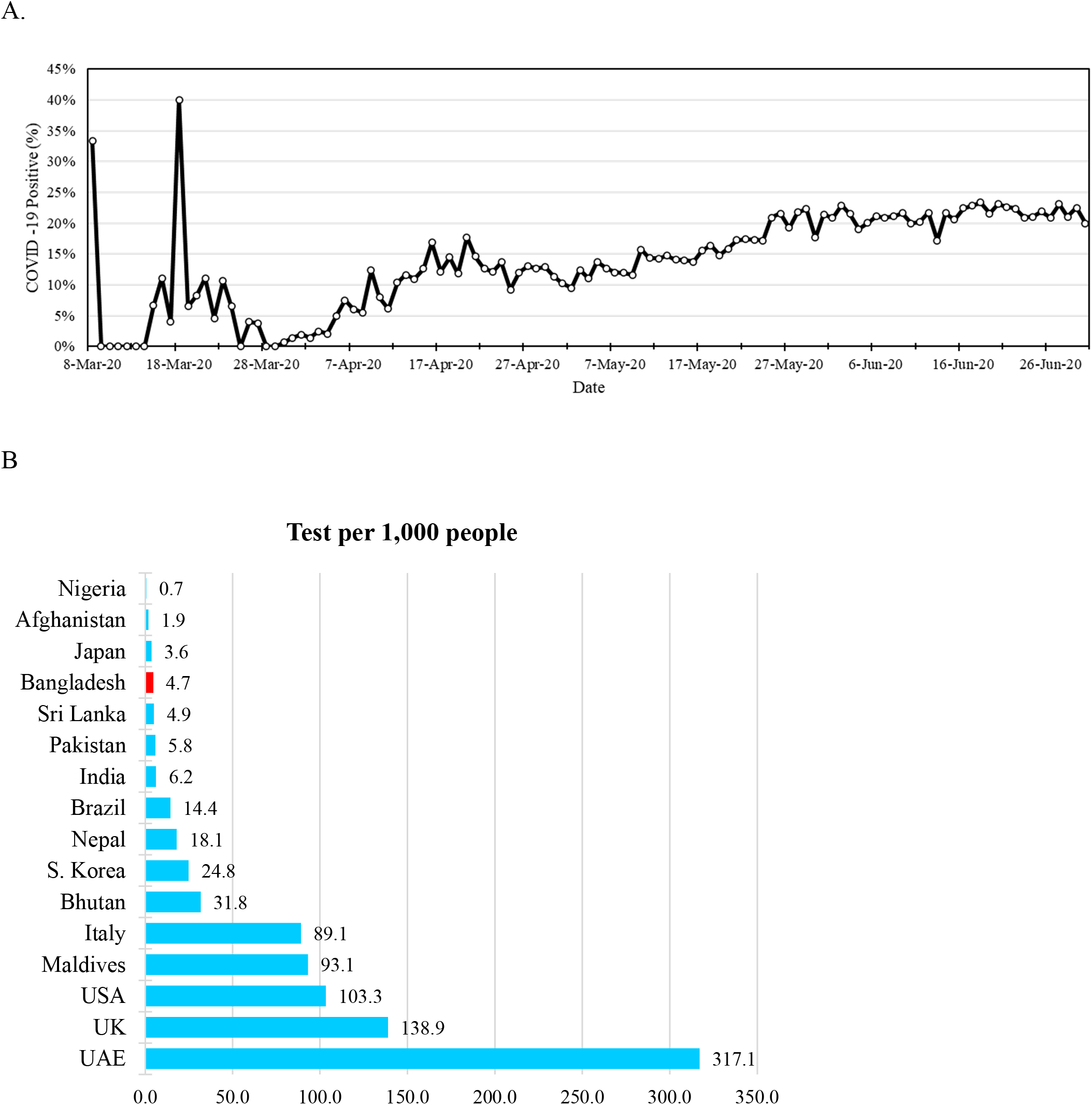
Overall scenario of RT-PCR based COVID-19 testing in Bangladesh. A) Percentage of positive cases of total test performed. B) Testing capacity of Bangladesh as compared to other countries.

## Notes

### Competing Interest Statement

The authors have declared no competing interest.

### Author Declarations

Not applicable since seconadry data were used

